# Patient-Reported Symptom Assessments and Early Chemotherapy Discontinuation in Routine Gastrointestinal Cancer Care

**DOI:** 10.1101/2025.10.07.25337194

**Authors:** Chengbo Zeng, Mariem Ahmed, Yu-Jen Chen, Shumenghui Zhai, Sandra C. Olisakwe, Patricia C. Dykes, Thomas J. Roberts, Nneka N. Ufere, Elizabeth S. Davis, Li Zhou, Maria O. Edelen, Andrea L. Pusic, Kelly M. Kenzik, Kelsey S. Lau-Min, Jason B. Liu

## Abstract

**Background:** Early chemotherapy (CTh) discontinuation in gastrointestinal (GI) cancer patients can compromise treatment effectiveness and worsen outcomes. Monitoring and addressing patient-reported symptoms may reduce early discontinuation. Yet, evidence baseline symptoms routinely collected in standard clinical practice remains limited.

**Methods:** This retrospective cohort study included adult patients with GI cancers who received CTh at Mass General Brigham between 01/2019 and 01/2024. The Patient-Reported Outcome version of the Common Terminology Criteria for Adverse Events assessed 12 symptoms at CTh initiation and at visits proximal to 30, 60, and 90 days from initiation. All patients completed baseline assessment. The primary outcome was early discontinuation due to toxicity within 90 days of initiation. Using the Fine-Gray model to account for competing risks of death and progression, we examined the association between early discontinuation and number of completed assessments over time. We tested whether this association differed across prognostic groups. To address time-varying bias from disease severity, treatment, and prior completion, we applied inverse probability weighting.

**Results:** Among 1,178 patients, the most common cancers were colorectal (35%) and pancreatic (31%), and 45% had stage IV disease. Overall, 784 (67%) completed assessments at CTh initiation and during follow-up. After adjusting for demographic and clinical covariates and time-varying bias, completing post-initiation assessments was associated with a lower cumulative risk of early discontinuation (SHR: 0.70 [95% CI: 0.44–1.12]). Results were generally consistent across prognostic groups.

**Conclusion:** More frequent completion of patient-reported symptom assessment post CTh initiation was associated with a lower risk of early discontinuation in practice.

## Introduction

Chemotherapy (CTh) remains central to the treatment for locally advanced and metastatic gastrointestinal (GI) cancers, yet adverse effects from CTh and symptoms related to the underlying malignancy can lead to premature treatment discontinuation and compromise patient quality of life and survival. In clinical trials, patient-reported symptom assessments can help identify adverse events, guide timely management, reduce early CTh discontinuation, and ultimately improve quality of life and survival.^1–5^ These assessments are monitored remotely by healthcare providers, nursing staff, and patients. Whether the benefits of symptom-assessment hold in routine care settings where the assessment is delivered before or during a clinic visit, however, remains underexplored. Specifically, evidence is needed to determine whether a dose-response relationship exists; that is, whether greater completion of symptom assessments is associated with a lower risk of early CTh discontinuation.

Implementing patient-reported symptom assessments in clinical practice is often more challenging than in clinical trials.^6,7^ Unlike remote symptom monitoring conducted as an intervention outside of clinical sites, routine collection of patient-reported symptoms usually involves diverse patient populations with varying demographic characteristics, cancer types, and disease stages and is typically integrated into regular clinical visits. Patients may complete symptom assessments synchronously or asynchronously with their visits, allowing the reported information to be used for symptom management and treatment plan adjustment, with the overall goal to improve care quality. However, large-scale implementation across diverse patient populations and within the healthcare systems is complex, and clinic-based patient-reported assessments are susceptible to nonresponse, which is difficult to track in routine care, leading to insufficient data to guide symptom management, prevent early discontinuation, and preserve outcomes.^8,9^ We hypothesized that greater completion of symptom assessments would be associated with a reduced risk of early CTh discontinuation, as more complete data would enable clinicians and patients to manage symptoms proactively.

Using routinely collected patient-reported symptom data from patients with GI cancers receiving CTh, we examined the association between symptom assessment completion and early CTh discontinuation. Specifically, we aimed to address two key research questions: (1) Is higher completion of symptom assessments associated with a lower risk of early CTh discontinuation? And (2) Does this association vary across prognostic groups, including age, degree of comorbidity, cancer stage, and site?

## Methods

### Study design, setting, and data source

This retrospective observational cohort study used routinely collected patient-reported symptom data and electronic health records (EHR) from the Mass General Brigham (MGB) health system. MGB is a large, integrated health system comprised of two academic medical centers, eight community hospitals, and 30 oncology practices. MGB has implemented the routine collection of patient-reported outcome measures (PROMs) for clinical care, including in medical oncology practices, wholly within its EHR. Details have been previously reported.^6,10^

At MGB GI cancer clinics, the symptom assessment program launched in 2019 is part of the standardized PROMs initiative, which collects the Patient-Reported Outcome (PRO) version of the Common Terminology Criteria for Adverse Events (PRO-CTCAE^®^) from all clinic visits. Patients with scheduled appointments are asked to complete the symptom assessment one week before their visit through the EHR patient portal. Those who do not complete the symptom assessment in advance are offered tablets to do so during their clinic visit. The symptom assessments are available in seven languages (i.e., English, Spanish, Portuguese, Haitian Creole, Traditional Chinese, Russian, Arabic). Reported scores are available immediately to clinicians in the EHR.

### Study population

We included adults between 01/2019 and 01/2024 who met the following criteria: (1) had a diagnosis of all-stage colorectal, esophageal, hepatobiliary, pancreatic, or gastric cancers based on ICD-10 codes (**Supplemental Table S1**), (2) initiated any cytotoxic chemotherapy (CTh) regimens with or without concurrent immunotherapy or targeted therapy to GI cancers, as determined by treatment regimens and their department specialties in the EHR, and (3) completed a patient-reported symptom assessment within 15 days of CTh initiation (i.e., the start of a new CTh episode), with possible additional completions at days 30, 60, and 90 post-initiation. During the study period, some patients underwent multiple episodes of CTh due to treatment adjustments. Because their clinical visits to GI cancer clinics could occur at varying times throughout their treatment courses, not all visits necessarily fell within the four pre-specified time windows (i.e., –15 to 14 [CTh initiation], 15 to 44 [day 30], 45 to 74 [day 60], and 75 to 104 [day 90] days). To ensure sufficient clinical visits with symptom assessments within the first 90 days after CTh initiation, we retained, for patients with multiple CTh episodes, the one that had the greatest number of clinical visits during this period. As a result, each patient contributed one CTh episode to the final analysis.

For patients who completed multiple assessments around the four pre-specified time windows, we only selected the one closest to each time point, resulting in a total of 1 to 4 completions per patient. Each completion yielded symptom data, so a patient had symptom data at CTh initiation and might also have data post initiation. **Supplemental Section A** provides a detailed description of the processing of routinely collected EHR and patient-reported symptom assessments.

### Patient-reported Symptoms

Details of the PRO-CTCAE surveys used here have been previously described.^11,12^ Patients were asked to report on the 12 most common CTh-related symptoms: constipation, decreased appetite, diarrhea, dyspnea, fatigue, fever, insomnia, nausea, paresthesia, pain, rash, and vomiting.^13–18^ For each symptom, patients are asked to report on its presence, frequency, severity, and/or interference. Assessment response for each symptom is assigned a composite score ranging from 0 to 3, where 0 represents no symptoms and 3 represents severe symptoms.^11,12^

Because we were interested in the relationship between number of patient-reported symptom assessment completion and the risk of early CTh discontinuation, the exposure of interest was the total number (ranging from 1 to 4) of assessments completed across the four pre-specified time points.

### Early discontinuation of Chemotherapy

We defined early CTh discontinuation as any deviations from the CTh treatment plan (e.g., dose reduction, regimen switches, and treatment stop) within 90 days of initiation.^19,20^ Reasons for discontinuation were obtained through independent manual chart review by three members (CZ, MA, and JL) of the research team and categorized into three groups: (1) toxicity, (2) disease progression, or (3) death. Treatments completed within 90 days were not considered discontinuation. Our primary analytic focus was on CTh discontinuation due to toxicity, as toxicity can be managed through timely and appropriate care from clinicians receiving the patient-reported symptom scores.

### Demographic and clinical characteristics

Demographic characteristics included age, sex, race/ethnicity, level of education, employment status, and insurance type. Clinical characteristics included National Cancer Institute Charlson comorbidity index (CCI: 0 or ≥1), primary cancer site, time since diagnosis, and cancer stage (i.e., I, II, & III vs. IV). Additional clinical details included the concurrent use of immunotherapy, radiation therapy, and targeted therapy during CTh (all coded as Yes/No), common CTh drugs (i.e., 5- fluorouracil, oxaliplatin, irinotecan, paclitaxel, and gemcitabine), and treatment goals (i.e., adjuvant/neoadjuvant, control, curative, maintenance/support, palliative).

### Statistical analysis

In the study cohort, we first summarized the number of completed symptom assessments across the four time points. We then compared baseline symptom burden across groups with different numbers of completed assessments using analysis of variance (ANOVA). Because the likelihood of completing an assessment may change over time due to factors such as disease severity, CTh-related adverse events, or prior completion behavior, this creates potential time-varying bias. To address this, we applied inverse probability (IP) weighting to the ANOVA and subsequent regression analyses.^21–23^ Details of the IP weight construction are provided in **Section B of the Supplemental Materials**.

We calculated the frequency and percentage of each reason for early discontinuation (i.e., toxicity, death, and disease progression). To address our first research question (“Is higher completion of symptom assessments associated with a lower risk of early CTh discontinuation?”), we used the Fine-Gray model to account for death and disease progression as competing risks. We further explored whether this association was consistent across age groups (< or ≥ 65 years), degree of comorbidity (CCI < or ≥ 1), cancer stage (i.e., I, II, & III vs. IV), and site (pancreatic cancer vs others), as these are the important factors associated with prognosis.^24–26^ This analysis helped address our second research question: does this association vary across prognostic groups? We reported sub distribution hazard ratios (SHR) and 95% confidence intervals (CI). We also compared these results with a standard Fine-Gray model with baseline covariates only. In these analyses, we adjusted for age, sex, race, level of education, cancer sites, cancer stage, concurrent target therapy, concurrent immunotherapy, concurrent radiation therapy, use of 5-fluorouracil, use of oxaliplatin, use of gemcitabine, comorbidity, and symptom score at CTh initiation, all of which were associated with the completion of symptom assessment **(Table S2)** and/or the risk of early discontinuation **(Table S3)** according to our bivariate analyses and existing literature.^27^

Lastly, we conducted secondary analyses to examine the association between discontinuation and symptom burden cross-sectionally using data at CTh initiation. We also examined their longitudinal association using data across the four time points.

Symptom burden was defined as the sum score of the 12 symptoms assessed. We employed multiple imputation with 50 replicates to handle missing symptom data. All statistical analyses were conducted using SAS version 9.4 (SAS Institute, Inc.), with a statistical significance level set at the two-sided p<0.05.

### Ethics Review

This study was approved by the Dana-Farber/Harvard Cancer Center (DF/HCC) Institutional Review Board (IRB protocol number: 25-048).

## Results

### Overview

Of the 1,178 eligible participants (**Figure 1**), more than half were older adults (52.4%), male (58.6%), White (86.1%), non-Hispanic (90.8%), and unemployed (56.9%) (**Table 1**). The most common cancers were colorectal (34.6%) and pancreatic (30.8%), and about 45% had stage IV cancers. The most frequently administered CTh drugs were 5-fluorouracil (65.5%), oxaliplatin (55.1%), and irinotecan (33.8%). About 45% of the patients received CTh with palliative intent.

**Figure 1.**
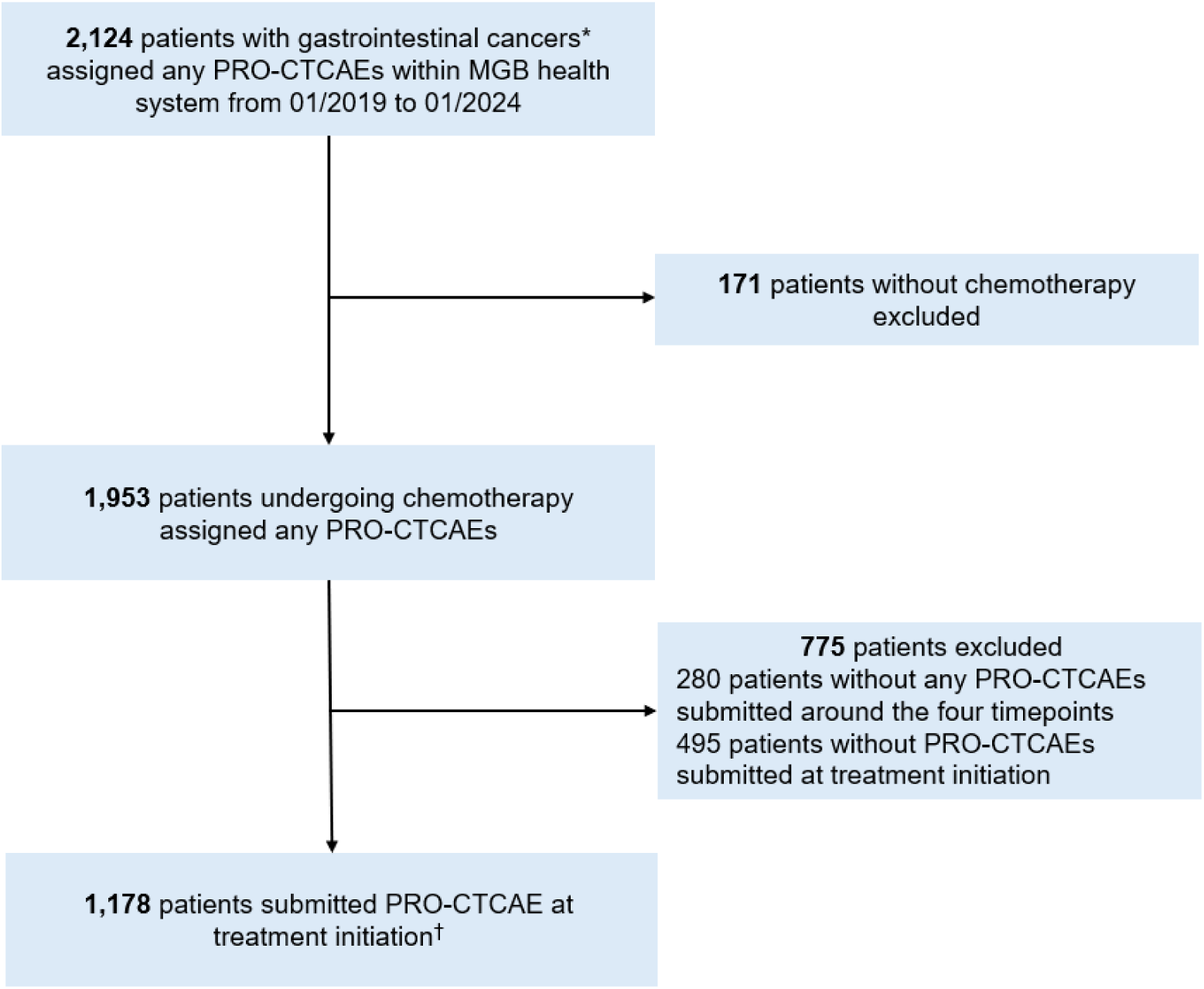
Inclusion and exclusion flowchart for patients with gastrointestinal cancers completing symptom assessment within the Mass General Brigham Health System, 2019-2024 *Notes:* *: any colorectal, esophagus, liver and bile duct, pancreatic, and stomach cancers. †: patients might also submit PRO-CTCAE at days 30, 60, and 90 post initiation.

**Table 1.**
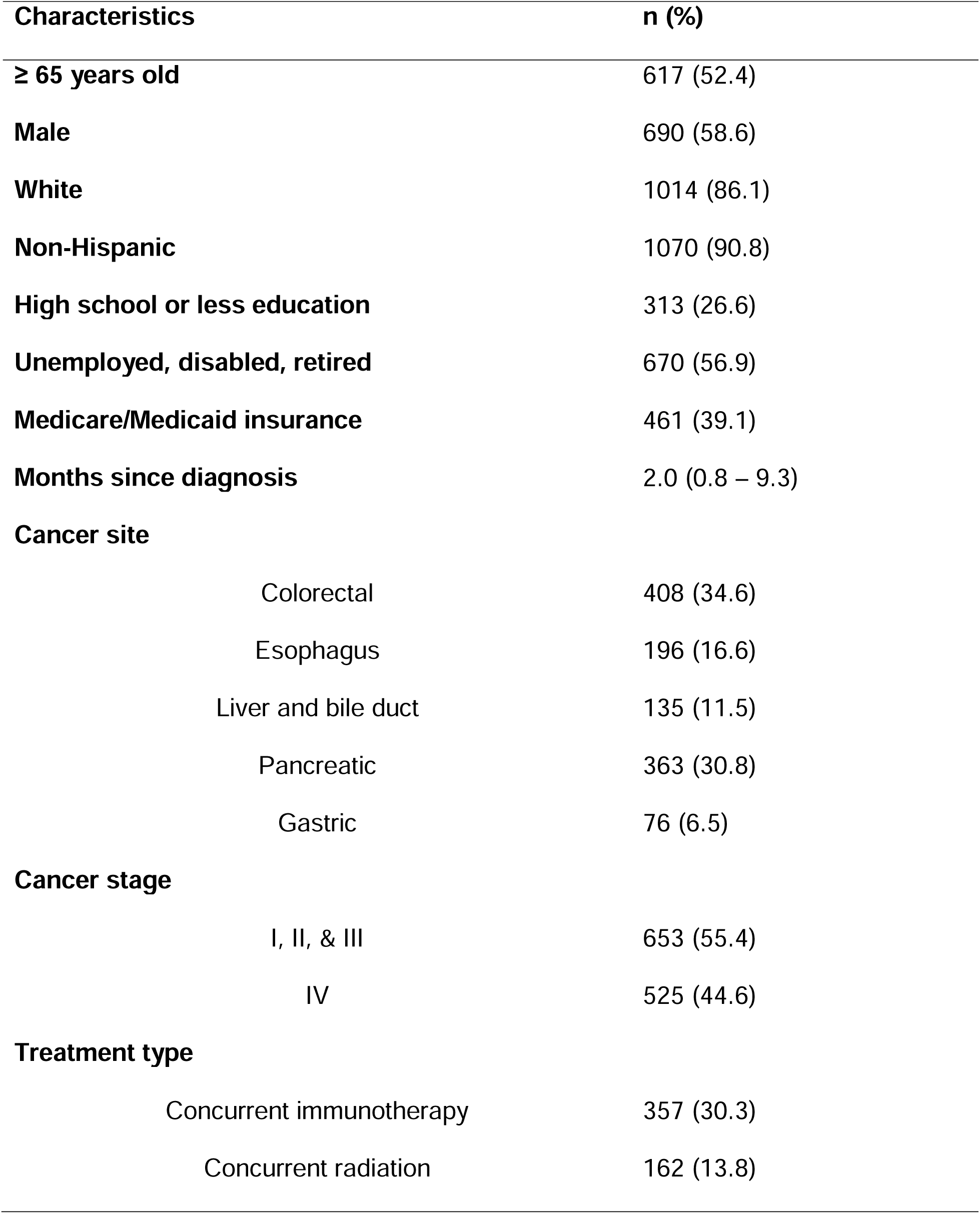
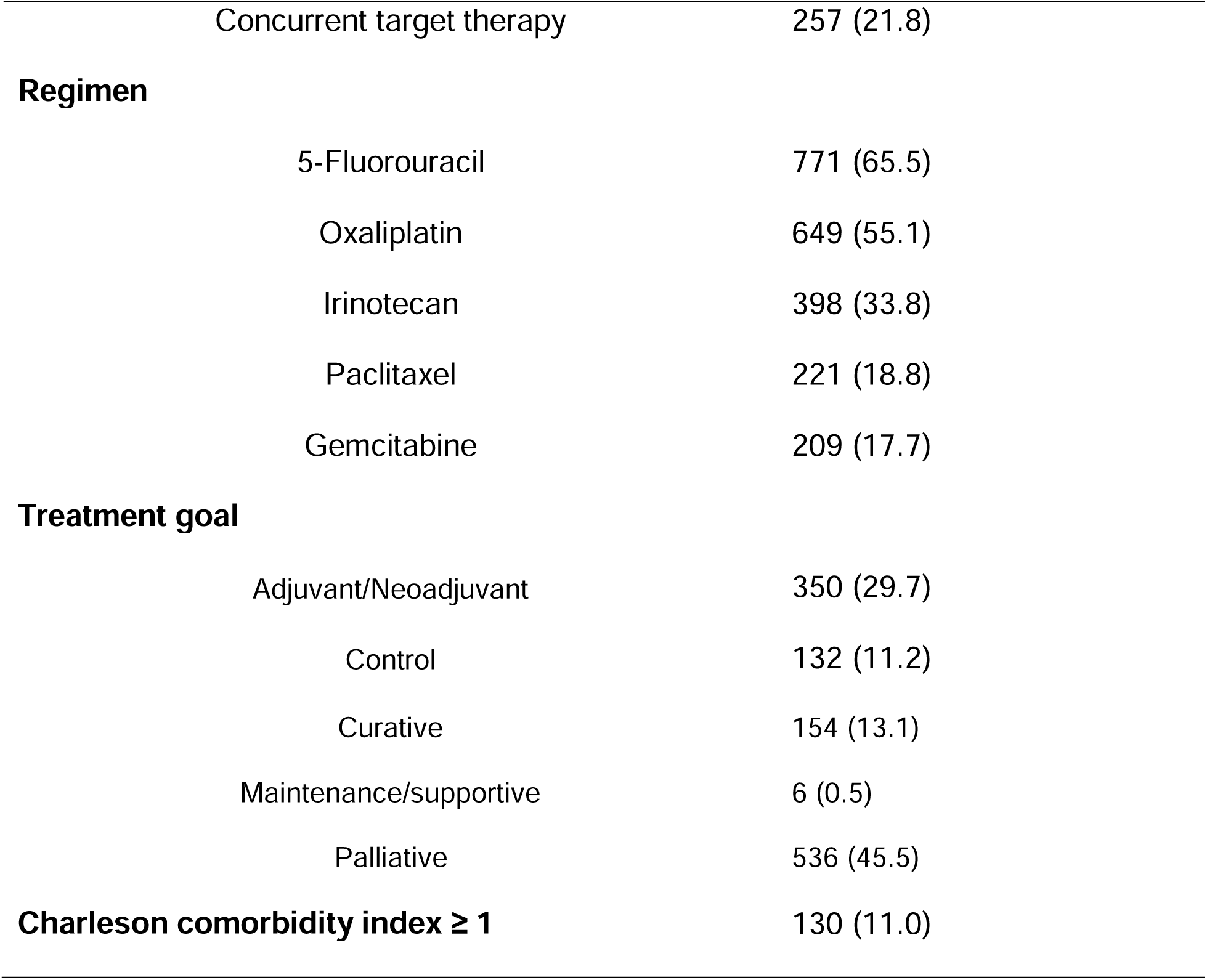
Demographic and clinical characteristics of patients with gastrointestinal cancers receiving chemotherapy, 2019-2024 (N=1,178)

### Distribution of symptom assessment completion and symptom burden

All patients completed assessments and had a clinical visit at CTh initiation, of whom 76.4% completed additional assessments and had subsequent visits at days 30, 60, or 90. In total, 16.2% completed three, and 26.2% completed all four. **Table 2** shows the distribution of symptom monitoring completions across the four time points and their combinations.

**Table 2.** Distribution of symptom monitoring completion in the study cohort across the four time points (N= 1,178)

| 1<sup>st</sup> completion | 2<sup>nd</sup> completion | 3<sup>rd</sup> completion | 4<sup>th</sup> completion | N | % |
| --- | --- | --- | --- | --- | --- |
| CTh initiation |  |  |  | 394 | 23.6 |
| CTh initiation | Day 30 |  |  | 211 | 12.6 |
| CTh initiation | Day 30 | Day 60 |  | 116 | 6.9 |
| CTh initiation | Day 30 | Day 60 | Day 90 | 309 | 18.5 |
| CTh initiation | Day 30 | Day 90 |  | 45 | 2.7 |
| CTh initiation | Day 60 |  |  | 48 | 2.9 |
| CTh initiation | Day 60 | Day 90 |  | 30 | 1.8 |
| CTh initiation | Day 90 |  |  | 25 | 1.5 |
| **Total** |  |  |  | **1,178** | **100** |
| One completion (Only CTh initiation) |  |  |  | 394 | 23.6 |
| Two completions (CTh initiation and one follow-up time point) |  |  |  | 284 | 24.0 |
| Three completions (CTh initiation and two follow-up time points) |  |  |  | 191 | 16.2 |
| Four completions (CTh initiation and three follow-up time points) |  |  |  | 309 | 26.2 |

Notes: CTh: chemotherapy

**Figure 2** displays mean symptom burden stratified by the number of symptom assessment completion at each time point. Among patients with only one symptom assessment completion (n=394), all contributed to the symptom score at CTh initiation, with a mean score of 8.4. For patients with additional assessments, the mean scores at CTh initiation were 8.1 for those with a total of two assessments (n=284), 7.4 for three (n=191), and 6.4 for four (n=309), respectively. There was significant difference among these mean symptom scores (p<0.001).

**Figure 2.**
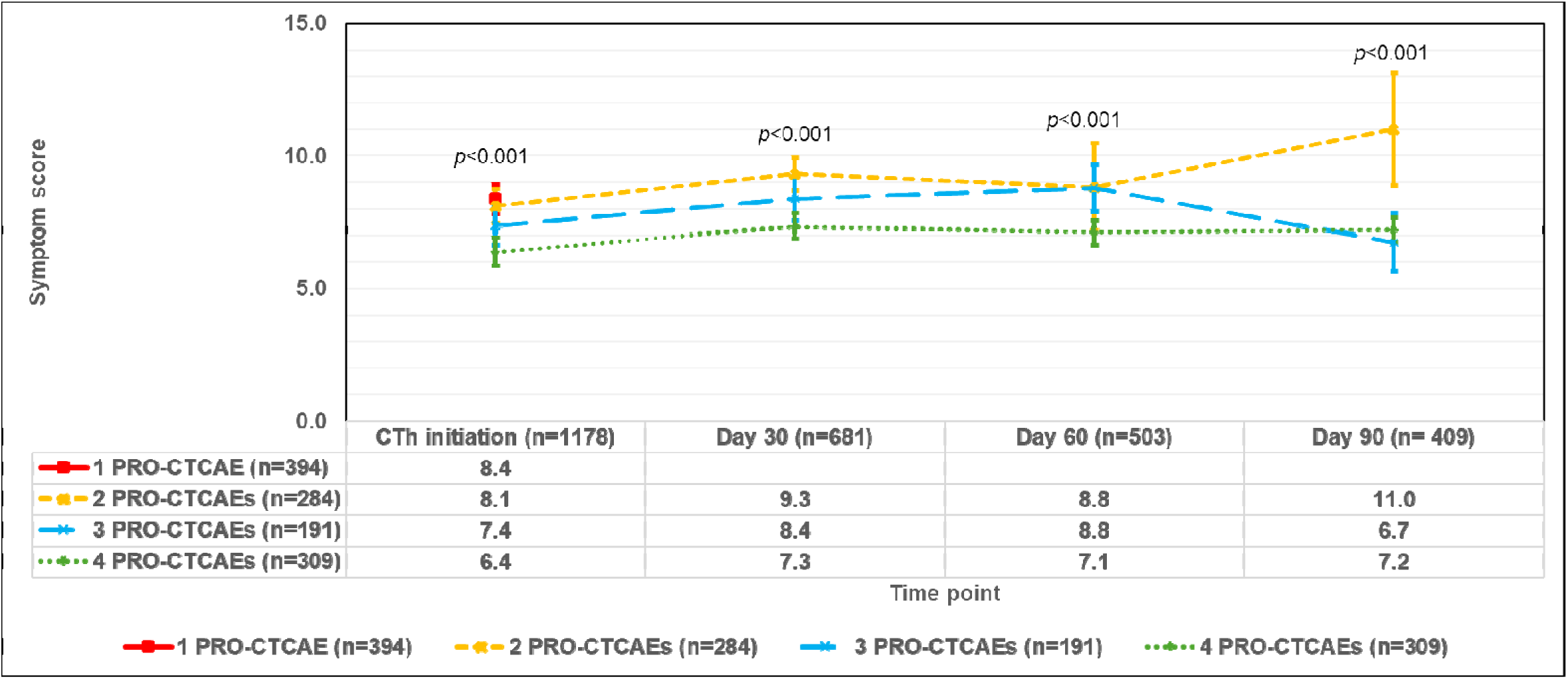
Symptom score by number of symptom assessments in patients with gastrointestinal cancers undergoing chemotherapy the Mass General Brigham Health System, 2019-2024 *Notes:* All patients with symptom assessments at chemotherapy (CTh) initiation were included. Patients with only one PRO-CTCAE contributed only to the symptom score at CTh initiation. Patients with at least two PRO-CTCAEs contributed to the symptom score at CTh initiation and at other time points (i.e., days 30, 60, and 90 post-initiation), depending on when they completed their symptom assessments.

Among patients with two symptom assessments, those with data at day 30 post CTh initiation had a mean score of 9.3; the mean was 8.4 for those with three assessments and 7.3 for those with four. We found significant differences in these scores (p<0.001). The mean scores among patients with data at days 60 and 90 are presented in **Figure 2**.

### Early discontinuation and number of symptom assessment completion

A total of 124 (10.5%, 124/1,178) patients discontinued CTh within 90 days. Among these, 19 (15.3%) discontinued due to toxicity, 47 (37.9%) due to death, and 58 (46.8%) due to disease progression (**Figure 3**). After accounting for the competing risks of death and disease progression and time-varying bias, the weighted model revealed that completing additional symptom assessments post CTh initiation was associated with a lower risk of early discontinuation (SHR = 0.70; 95% CI: 0.44–1.12; **Table 3**), although this association was not significant. Subsequent analyses across age groups, degree of comorbidity, cancer stage, and site revealed similar findings.

**Figure 3.**
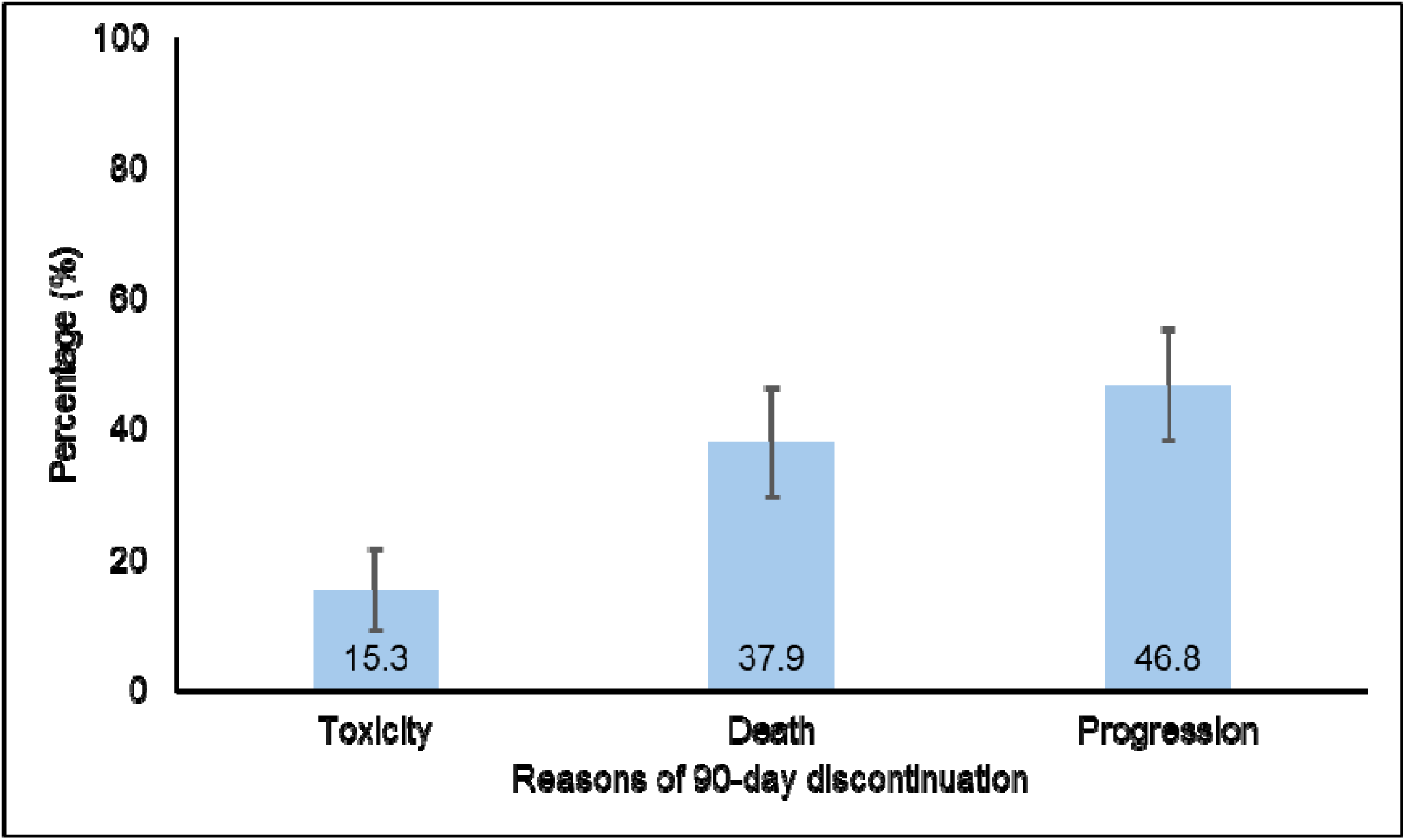
Reasons and distributions of 90-day discontinuation in patients with gastrointestinal cancers receiving chemotherapy, 2019-2024 (N=124)

**Table 3.** Fine-Gray model of 90-day discontinuation due to toxicity predicted by the number of symptom assessment completion in patients with gastrointestinal cancers receiving chemotherapy, 2019-2024

|  | Subdistribution hazard ratio (95%CI) |  |
| --- | --- | --- |
|  | Unweighted | Weighted |
| **Overall sample** |  |  |
| **Number of completion** | **0.49 (0.25 – 0.95)** | 0.70 (0.44 – 1.12) |
| **Age groups** |  |  |
| ≤ 64 years | 0.72 (0.42 – 1.24) | 0.99 (0.64 – 1.52) |
| > 64 years | **0.27 (0.07 – 0.99)** | 0.31 (0.08 – 1.17) |
| **Comorbidity** |  |  |
| CCI = 0 | 0.62 (0.37 – 1.06) | 0.80 (0.50 – 1.27) |
| CCI ≥ 1* | - | - |
| **Cancer stage** |  |  |
| I, II, & III | 0.63 (0.34 – 1.17) | 0.85 (0.49 – 1.47) |
| IV | 0.33 (0.10 – 1.10) | 0.58 (0.31 – 1.06) |
| **Cancer site** |  |  |
| Pancreatic cancer | 0.50 (0.17 – 1.43) | 0.79 (0.41 – 1.55) |
| Other cancers | **0.48 (0.24 – 0.97)** | 0.65 (0.36 – 1.15) |

*Notes:* We controlled age, sex, race, level of education, cancer site, cancer stage, concurrent immunotherapy, concurrent radiation therapy, use of 5-fluorouracil, use of oxaliplatin, use of gemcitabine, comorbidity, symptom score at treatment initiation. CCI: Charlson comorbidity index. \*: Model was not converged due to small sample size.

The unweighted model without adjusting for time-varying bias found significant relationship between number of symptom assessment completion and early discontinuation. Subsequent analyses across age group, degree of comorbidity, cancer stage, and site found consistent trends, but we only found statistical significance in older adults and patients without pancreatic cancer.

### Secondary analyses of early discontinuation and symptom burden

Our secondary analyses found that whether using symptom burden at CTh initiation or its change during the first 90 days of CTh, higher symptom burden was associated with an increased risk of discontinuation due to toxicity, although the association was only marginally significant **(Table S4)**.

## Discussion

In these data, we observed a trend suggesting more frequent patient-reported symptom assessment completion, along with clinical visits during CTh, was associated with a lower cumulative risk of early discontinuation due to toxicity; however, this relationship was not statistically significant. Notably, over 60% of the patients had assessments at both CTh initiation and post-initiation.

High-certainty evidence from clinical trials supports the use of patient-reported symptom assessment to reduce adverse patient outcomes;^4,5^ however, the generalizability of these findings to routine clinical practice remains unclear.^28,29^ Our study intended to bridge this knowledge gap by examining the relationship between the number of patient-reported symptom assessments completed in conjunction with routine clinical visits and the risk of early discontinuation due to toxicity. Unexpectedly, after adjusting for the time-varying bias and competing events, this association was not statistically significant; however, we detected a directional trend toward reduced discontinuation with more assessments, and thus more clinical visits, being completed. As an observational study, our findings serve to generate hypotheses for future study.

Our findings may be partially explained by the low incidence (10.5%) of early discontinuation due to toxicity within 90 days of CTh initiation. Another possibility is that patient-reported symptom data may not be used in real time by clinicians for treatment planning. Unlike in controlled clinical trial settings, the use of patient-reported symptom data in clinical care is often influenced by clinicians’ workload and by how both clinicians and patients perceive the utility of these activities and data. A large mixed-methods study found that over 20% of clinicians did not review patient-reported data with their patients, and 44% regarded the data as unhelpful or held a neutral attitude toward its usefulness.^30^ This underscores the pervasive research-to-practice phenomenon that exists across clinical research, not only in the realm of patient-reported symptom monitoring.

Although not statistically significant, our findings have important implications for implementing patient-reported symptom assessment in routine cancer care. We found patients who completed symptom assessment at CTh initiation were found to have a lower risk of discontinuation if they also complete symptom assessment during their subsequent clinical visits. Conducting symptom monitoring in routine care during CTh can be challenging, as higher toxicity and lower patient engagement are often associated with lower response rates.^6,27^ While it is difficult to ensure completion of symptom assessment throughout the entire treatment course, our findings suggest that collecting patient-reported symptoms at CTh initiation and at least one follow-up time point may be a practical and effective strategy. This approach provides clinicians and patients with symptom data both before and after CTh initiation, allowing for the tracking of progress and the detection for clinically meaningful changes. It may support better management of CTh-related side effects and potentially reduce early discontinuation within the first 90 days.

### Strengths

Our study leveraged a real-world patient cohort from routine GI cancer clinics within an integrated health system that has a standardized, well-established PROMs program. We focused on the number of symptom assessment completion and examined its dose-response relationship with the risk of early discontinuation among patients who were asked to report symptoms during their CTh. Our analytic approach was carefully designed to minimize time-varying bias inherent in observational EHR data. For example, recognizing that patients’ ability and willingness to complete symptom assessment change over time, we applied inverse probability weighting to adjust for time-varying bias introduced by disease severity, treatment, and prior completion. We also employed the Fine-Gray model to account for competing risks, such as discontinuation due to death and disease progression. These methodological strategies, which are applicable to research involving routinely collected longitudinal PRO data, enhance the internal validity of our findings and support the generation of high-quality, real-world evidence with strong clinical relevance and implications for routine cancer care.

### Limitations

Our findings should be interpreted in the context of several limitations. First, despite the study’s rigorous design and analytic approach, it was observational in nature and cannot establish a causal direction between symptom assessment and early CTh discontinuation. However, existing clinical trial evidence has shown that assessing and addressing patient-reported symptoms results in improved clinical outcomes. Thus, we aimed to provide complementary evidence from routine care to examine how clinical trial learnings support symptom assessment and management in practice. Second, the relatively low incidence of 90-day discontinuation due to toxicity limited our ability to detect significance in statistical inference. Future studies with a larger sample size and longer timeframe are needed to validate these results. Third, while IP weighting helps reduce bias across patients with varying number of symptom assessment completion at different time points, it relies on the assumption that all relevant confounding variables were included in generating the weights. Due to limitations in data availability, our analysis may have omitted other important confounders. Finally, this study was conducted within a single healthcare system in the Boston metropolitan area. Therefore, caution is needed when generalizing these findings to other health systems or regions of the country.

## Conclusions

Our findings suggest a trend toward more frequent completion of patient-reported symptom assessments along with routine clinical visits being associated with a lower risk of early CTh discontinuation. Integrating patient-reported symptom assessment into routine care and using these data to guide medical decision-making represent a critical next step towards improving treatment safety and supporting the delivery of high-quality care during CTh.

Statement and Declarations

## Data Availability

Individual patient data will not be shared.

## Acknowledgments

The authors thank the clinicians and program staff for their support on the successful implementation of patient-reported outcome measure collection at the gastrointestinal cancer clinics. The abstract related to this work has been published in the JCO Oncology Practice. Chengbo Zeng et al. Patient-reported symptom assessments and premature chemotherapy discontinuation in patients with gastrointestinal cancers. JCO Oncol Pract 21, 383-383(2025). DOI:10.1200/OP.2025.21.10_suppl.383

## Data availability statement

Individual patient data will not be shared.

## Ethical approval statement and consent to participate

This study was approved by the Dana-Farber/Harvard Cancer Center (DF/HCC) Institutional Review Board (IRB protocol number: 25-048). This study involves secondary data analysis and does not require patient consent.

## **Section A.** Processing of routinely collected electronic health records and patient-reported symptom assessments

### Challenges of processing routinely collected data

Processing routinely collected electronic health records (EHR) and patient-reported symptom assessments poses substantial challenges. Patients often have multiple clinical visits during the study period, and the intervals between visits vary widely across individuals. Although patients with scheduled appointments are asked to complete a symptom assessment one week before their clinical visits through the standardized patient-reported outcome measures (PROMs) program at Mass General Brigham (MGB), not all patients complete these assessments. To enable rigorous analyses, we anchored clinical visits to key time chemotherapy (CTh) points and organized the data into a structured format aligned with our research objectives.

### Study population

The MGB gastrointestinal (GI) cancer clinics began implementing patient-reported symptom assessments through the MGB PROMs program in January 2019; therefore, our data extraction began at that time. Between January 2019 and January 2024, we identified adult patients with confirmed diagnoses of all-stage colorectal, esophageal, hepatobiliary, pancreatic, or gastric cancers based on primary diagnoses and corresponding ICD-10 codes **(Supplemental Table S1)**. We then identified a subset of patients who initiated any cytotoxic CTh regimens with or without concurrent immunotherapy or targeted therapy to GI cancers, as determined by treatment regimens and their department specialties in the EHR. Patients who underwent immunotherapy or targeted therapy only were excluded. Each included patient had at least one episode of CTh, and some had multiple episodes due to treatment adjustments during the study period.

### Mapping clinical visits and their associated symptom assessments to key chemotherapy time points

Our primary focus was patient-reported symptom assessments, and the main outcome was early CTh discontinuation within 90 days of initiation. Patients could have multiple clinical visits (e.g., 0, 1, 2, or more) to GI cancer clinics during CTh treatment. To ensure consistent alignment across patients, we mapped clinical visits to key CTh time points, which allowed us to maximize the use of available symptom assessments and completions in the analysis.

We pre-specified four key time points: CTh initiation and days 30, 60, and 90 post-initiation. Eligible CTh episodes were required to include at least one clinical visit occurring within 15 days of any of these time points, corresponding to four time windows: –15 to 14 (CTh initiation), 15 to 44 (day 30), 45 to 74 (day 60), and 75 to 104 (day 90) days. For patients with multiple CTh episodes, we retained the episode with the greatest number of clinical visits within these windows, resulting in one CTh episode per patient in the final analysis. For patients with multiple symptom assessments within the same window, we selected the one closest to the time point, resulting in one to four total assessments per patient. Finally, to ensure baseline completeness, only patients who completed a symptom assessment within 15 days of CTh initiation were included, with possible additional completions at days 30, 60, and 90 after initiation.

## Section B. Construction of inverse probability weight

The probability of completion is calculated at each time point. At chemotherapy initiation, the probability of completion (*p*_0_) is 1. For any subsequent time point *i*, the overall probability of completion (*P_i_*_;_) is the cumulative product of the conditional probabilities of completion from time 0 through time *i*, i.e. *p*_0_, …, *P_t_* The formula for calculating Pi isshown in Formula 1:

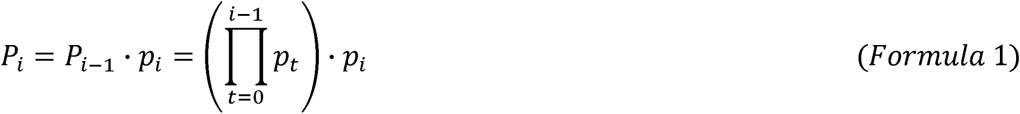

For each time point, the weight (*W_i_*) is calculated as the inverse of the probability of completion (*p_i_*) (Formula 2). The final weight was retained for each individual and adjusted in the analysis

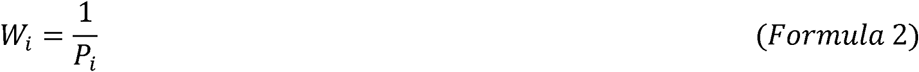

The probability of completion at each time point post chemotherapy initiation was modeled using age, sex, race, level of education, cancer site, cancer stage, concurrent immunotherapy, concurrent radiation therapy, use of 5-fluorouracil, use of oxaliplatin, use of gemcitabine, comorbidity, symptom score at CTh initiation, and prior completion.

---

**Table S1.** Cancer types and ICD-10 codes

| **Cancer Types** | **ICD-10 codes** |
| --- | --- |
| Colorectal | C18.X, C19, C20 |
| Esophagus | C15.X |
| Liver and bile duct | C22.X, C23 |
| Pancreatic | C25.X |
| Gastric | C16.X |

**Table S2.** Bivariate analysis of number of assessment completion (1 vs. ≥2) and demographic and clinical characteristics

| **Characteristics** | **p-value** |
| --- | --- |
| Age group | 0.866 |
| **Sex** | **0.056** |
| Race | 0.265 |
| Ethnicity | 0.320 |
| Education | 0.360 |
| Employment | 0.506 |
| Insurance type | 0.259 |
| Duration of diagnosis | 0.355 |
| **Cancer site** | **0.069** |
| Cancer stage | 0.196 |
| Concurrent immunotherapy | 0.629 |
| **Concurrent radiation** | **0.052** |
| Concurrent target therapy | 0.886 |
| **5-Fluorouracil** | **<0.001** |
| **Oxaliplatin** | **0.009** |
| Irinotecan | 0.504 |
| Paclitaxel | 0.111 |
| Gemcitabine | 0.251 |
| Charleson comorbidity index | 0.488 |
| **Symptom score at chemotherapy initiation** | **<0.001** |

*Notes:* Variables in bold were included in the construction of inverse probability weight.

**Table S3.**
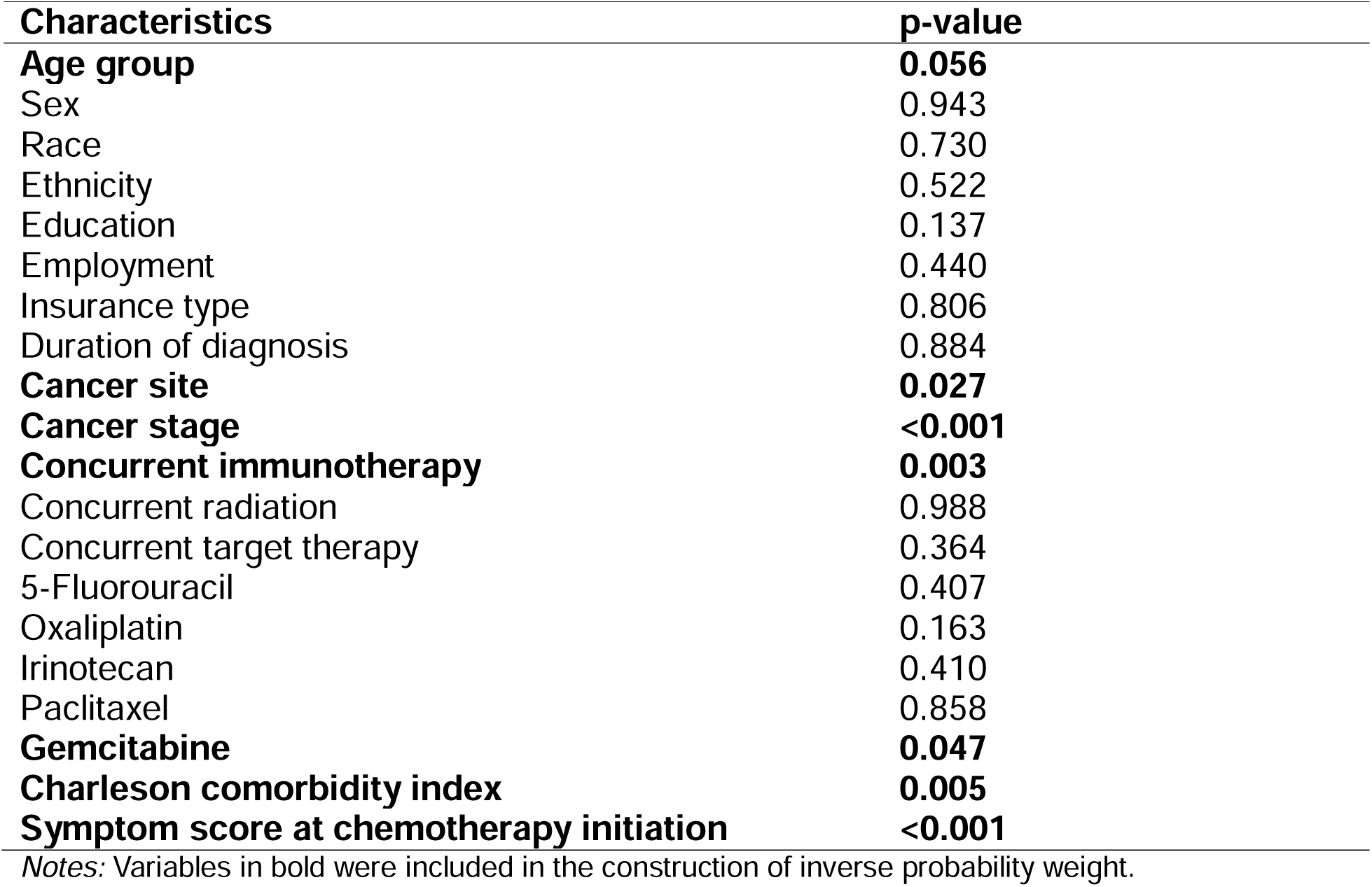
Bivariate analysis of discontinuation within the first 90 days of chemotherapy and demographic and clinical characteristics

**Table S4.** Fine-Gray model of 90-day discontinuation due to toxicity predicted by the symptom score in patients with gastrointestinal cancers receiving chemotherapy, 2019-2024^†^

|  | SHR (95%CI) |
| --- | --- |
| **Model 1** |  |
| Symptom score at CTh initiation | 1.03 (0.95 – 1.11) |
| **Model 2<sup>†</sup>** |  |
| Change in symptom score from CTh initiation to day 90 | 1.07 (0.97 – 1.17) |

*Notes:* †: Multiple imputation was used to handle missing data. SHR: Subdistribution hazard ratio. For these analyses, we controlled age, race, education, cancer site, concurrent target therapy, concurrent immunotherapy, concurrent radiation therapy, use of 5-FU, use of oxaliplatin, duration after diagnosis, comorbidity, and cancer stage.

